# A Personalized Probabilistic Approach to Ovarian Cancer Diagnostics

**DOI:** 10.1101/2023.11.24.23298971

**Authors:** Dongjo Ban, Stephen N. Housley, Lilya V. Matyunina, L. DeEtte McDonald, Victoria L. Bae-Jump, Benedict B. Benigno, Jeffrey Skolnick, John F. McDonald

**Affiliations:** Integrated Cancer Research Center, School of Biological Sciences, Georgia Institute of Technology, 315 Ferst Drive, Atlanta, GA 30332 USA; Department of Obstetrics and Gynecology, University of North Carolina, 3009 Old Clinic Building, Chapel Hill, NC 27599, USA; Ovarian Cancer Institute, 1266 W. Paces Ferry Rd NW #339, Atlanta, GA 30327, USA; Center for the Study of Systems Biology, School of Biological Sciences, Georgia Institute of Technology, 315 Ferst Drive, Atlanta, GA 30332, USA

## Abstract

**Objective:** The identification/development of a machine learning (ML)-based classifier that utilizes metabolic profiles of serum samples to accurately identify individuals with ovarian cancer (OC).

**Methods:** Serum samples collected from 431 OC patients and 133 normal women at four geographic locations were analyzed by mass spectrometry. Reliable metabolites were identified using recursive feature elimination (RFE) coupled with repeated cross-validation (CV) and used to develop a consensus classifier able to distinguish cancer from non-cancer. The probabilities assigned to individuals by the model were used to create a clinical tool that assigns a likelihood that an individual patient sample is cancer or normal.

**Results:** Our consensus classification model is able to distinguish cancer from control samples with 93% accuracy. The frequency distribution of individual patient scores was used to develop a clinical tool that assigns a likelihood that an individual patient does or does not have cancer.

**Conclusions:** An integrative approach using metabolomic profiles and ML-based classifiers has been employed to develop a clinical tool that assigns a probability that an individual patient does or does not have OC. This personalized/probabilistic approach to cancer diagnostics is more clinically informative and accurate than traditional binary (yes/no) tests and represents a promising new direction in the early detection of OC.

**HIGHLIGHTS:**

- Predictive models derived from machine learning (ML) analyses of serum metabolic profiles can accurately (PPV 93%) detect ovarian cancer (OC).
- Only a minority of the most predictively informative metabolites are currently annotated (7%).
- Lipids predominate among the most predictively informative metabolites currently annotated.
- The frequency distribution of model-derived patient scores can be used to develop a useful clinical tool for the diagnosis of OC.

## 1. Introduction

Early cancer diagnosis is one of the most important contributing factors to the successful treatment of the disease [1]. Early diagnosis is especially challenging for cancers like ovarian cancer (OC) that can progress rapidly, and yet display little to no clinical symptoms early in their development [2]. The ideal cancer diagnostic should not only be highly accurate, but additionally non-invasive and low cost to be widely available to the general public. Despite heroic efforts to develop such cancer diagnostics over the last several decades, this goal has proven to be frustratingly elusive [3]. A major reason for this is that, on the molecular level, cancer is a highly heterogenous disease not only between different types of cancer but even among individuals with the same cancer type [4]. As a consequence, finding a single molecular biomarker or set of biomarkers that are universally shared among individuals with even the same type of cancer is extremely difficult.

In recent years, various computational methods, including machine learning (ML), have been applied in efforts to identify patterns embedded within large omics datasets (*e.g*., genomic/proteomic/metabolomic) that may constitute an accurate diagnostic of cancer [5], [6] and other diseases [7]. For example, perturbations of metabolic levels in the blood and/or other body fluids have long been considered promising indicators of cancer and other diseases [8], [9] because metabolites constitute end points of many, if not most, of the molecular processes underlying biological functions. As such, metabolic profiles have been proposed as a molecular phenotype of biological systems, reflective of collective information encoded at the genome level and realized at the transcriptome and proteome levels [10].

Despite the inherent advantages of metabolic patterns as biomarkers of cancer and other diseases, extreme care is required in both the selection and analysis of metabolomic datasets. For example, potential technical inconsistencies in data acquisition (*e.g*., variation in sensitivity between instruments/laboratories and/or analytic drift associated with the same instrument over time) can easily compromise the reliability of acquired datasets unless frequent standardization with control samples is employed throughout the analytic process. In addition, extra precaution is needed in both the computational analysis of metabolic and other omics datasets and the interpretation of results. For example, there are a variety of ML approaches to the analysis of omics data, and each is associated with individual strengths and weaknesses [11], [12]. Despite these challenges, the use of metabolomic and other omics profiles as early indicators of cancer is not insurmountable and may provide clinicians with a powerful and highly accurate tool for personalized cancer diagnosis when properly addressed.

We report here on the development of a ML-based approach for the early detection of OC using metabolomic profiles in blood. Analyses were carried out on serum samples collected from 431 OC patients and 133 normal women at four geographic locations in the United States and Canada. The utility of a consensus classifier was evaluated using four independent sets of metabolomic profiles. Combining the best predictions from each profile using the consensus classifier resulted in a final set of predictions that can distinguish cancer from control samples with high accuracy (PPV 93%). We illustrate how the frequency distribution of individual patient scores can be used to develop a useful clinical tool that may be used to assign a likelihood that an individual does or does not have OC.

## 2. Methods

Details of the extensive methods employed in this study are presented in the Supplementary Material. Briefly, 431 serous papillary OC and 133 normal serum samples were obtained from four geographic locations (Atlanta, GA, Philadelphia, PA, Chapel Hill, NC, and Alberta, Canada) and were transferred to *Creative Proteomics* laboratory (Shirley, NY) for ultra-performance liquid chromatography, high-resolution mass spectrometry (UPLC-MS) analysis. A pooled quality control sample was obtained by combing equal amounts of each of the individual OC and control serum samples. Samples were individually processed through two different columns and analyzed using two different ionization modes (negative and positive) resulting in four distinct datasets (HP: HILIC positive; HN: HILIC negative; RN: C_18_ reversed phase negative; RP: C_18_ reversed phase positive). Reliable features (metabolites) were identified using recursive feature elimination (RFE) coupled with repeated cross-validation (CV). The output from these processing steps for each of the four datasets was an assignment of a relative ranking of features reflective of the relative frequencies of the features after repeated CV iterations. A consensus classifier was constructed by aggregating the results of five independent ML classifiers [logistic regression (LRC), random forest (RFC), support vector machine (SVM), k-nearest neighbor (KNN), and adaptive boosting (ADA)] to generate predictive classification models. The probabilities assigned to individuals by the consensus model were utilized to create a background distribution of probabilities that a given sample was cancer or normal.

## 3. Results

### 3.1 Data acquisition

The data acquisition process for this study is summarized in Figure 1A. Serum samples collected from 431 OC patients and 133 non-cancerous/normal individuals were obtained from four geographic locations in the United States (Fox Chase Cancer Center-Philadelphia, PA; UNC-Chapel Hill, NC; Northside Hospital-Atlanta, GA, and Canada-Alberta Health Services-Alberta, BC). Samples were characterized using ultra-performance liquid chromatography coupled with tandem mass spectrometry (UPLC-MS/MS). Each serum sample was independently processed through two different columns (HILIC and C_18_ reversed phase) and analyzed using two different ionization modes (negative and positive) resulting in four distinct datasets during MS/MS (HP: HILIC positive; HN: HILIC negative; RN: C_18_ reversed phase negative; RP: C_18_ reversed phase positive). Because of the large number of samples, metabolomic analyses were conducted over two separate batches. To detect and correct instrument drift within and between runs, a pooled quality control (QC) sample was run following analysis of every ten patient samples. A scatter plot of principle component analyses performed on the preprocessed data confirmed that no significant experimental variation was detected between batches after quality control of the data (Fig. 1B).

**Fig. 1.**
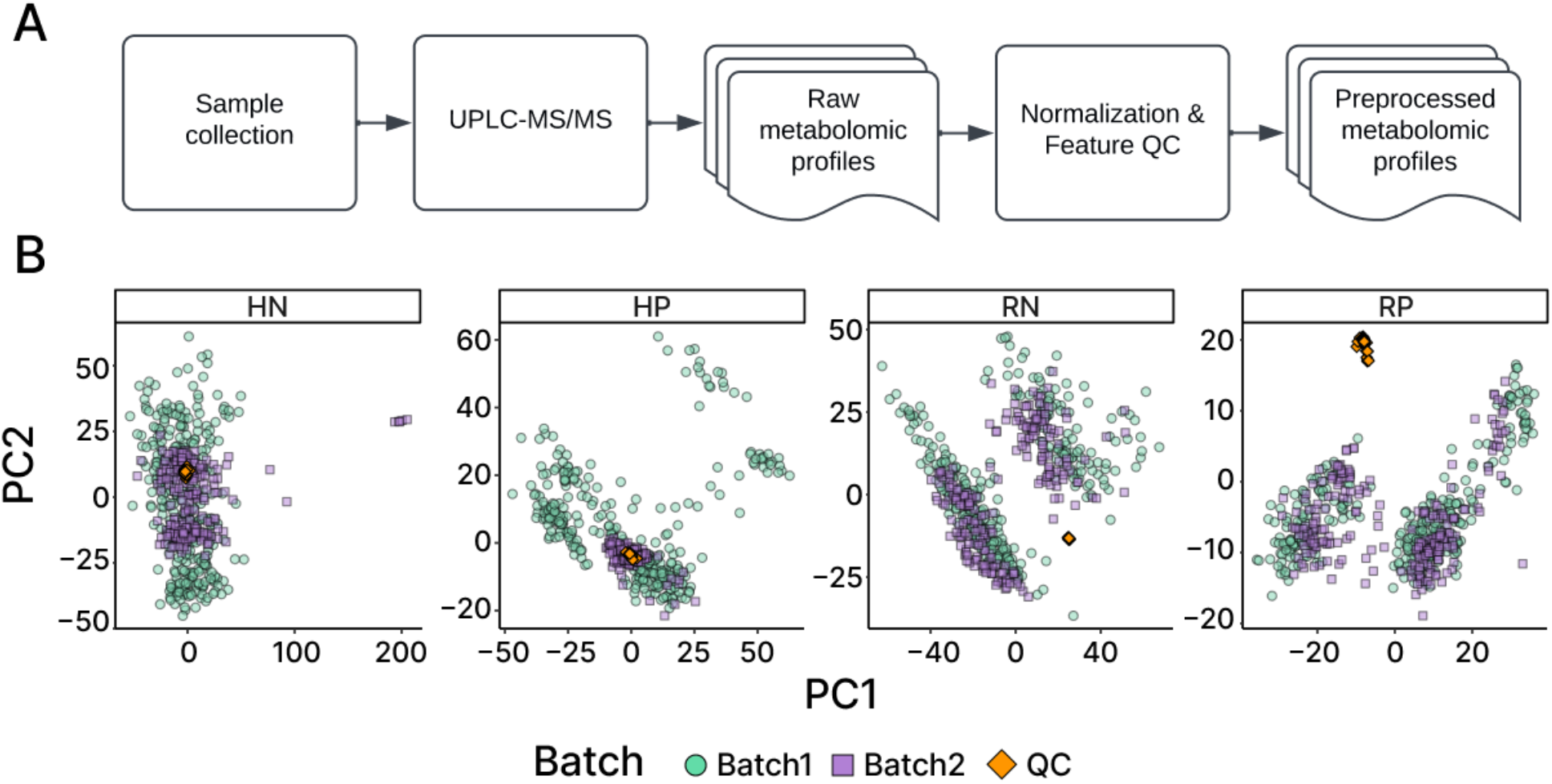
Workflow diagram illustrating data acquisition and preparation process. A) Serum samples from ovarian cancer patients and non-cancerous individuals are collected from multiple geolocations. They are analyzed using UPLC-MS/MS in an untargeted workflow to characterize the metabolome of ovarian cancer patients. Normalization and filtering of the features are performed following the best practices to obtain the preprocessed metabolomic profiles for downstream analyses. B) Scatter plot of principal component analysis performed on the preprocessed data after accounting for systematic and random errors. QC samples (orange) are shown to mostly cluster together with no clear separation between the two batches, indicating unwanted experimental variation has been eliminated.

### 3.2 Assessing the Stability of Metabolomic Features

The overall goal of our study is the identification/development of a ML classifier that utilizes metabolic profiles to accurately distinguish individuals with or without OC. Toward this end, we independently examined the predictive accuracy of five ML classifiers for each of the four datasets: RFC, SVC, ADA, KNN, and LRC.

Prior to the independent evaluation of each of these classifiers, we identified reliable features (metabolites) with recursive feature elimination (RFE) [13] coupled with repeated cross-validation (CV). The output from these processing steps for each of the four datasets was an assignment of a relative ranking of features reflective of the relative frequencies of the features after repeated CV iterations, as well as their relative contribution levels as determined by the Gini importance scores (see Methods in Supplementary Material for details).

Across all four datasets, we observed a moderate positive correlation (HN: R = 0.26, p < 0.001; HP: R = 0.56, p < 0.001; RN: R = 0.39, p < 0.001; RP: R = 0.50, p < 0.001) between the relative frequency of features and their importance (Fig. 2). This trend is most apparent in the RP dataset where the vast majority of features of high importance were in high frequency. In contrast, the HP dataset displayed a number of lower frequency features of high Gini importance (See Methods in Supplementary Material for details).

**Fig. 2.**
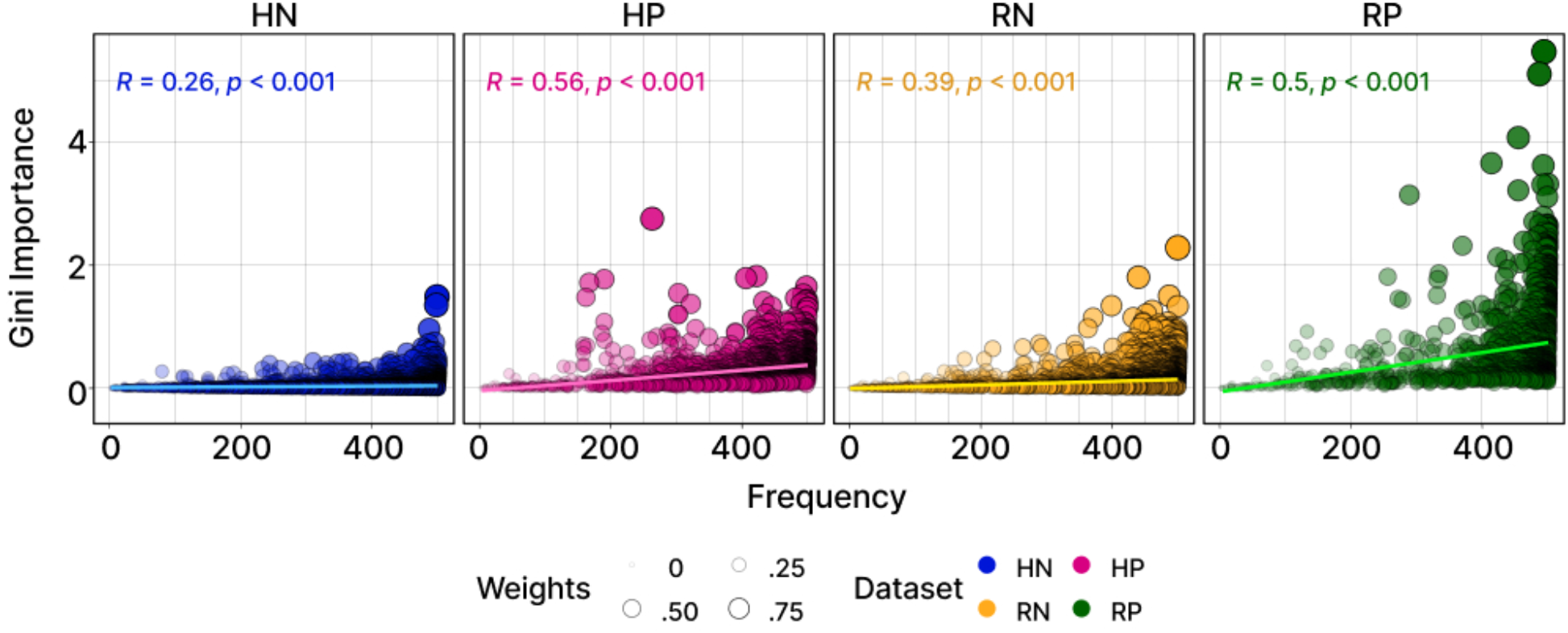
The frequency and Gini importance values of features in each dataset. The x-axis and y-axis correspond to feature frequency and importance value, respectively. The weights computed by combining frequencies and importance values were represented by the sizes and opacity of the points. The analysis revealed that across all datasets, many features had high frequency but relatively lower levels of importance. The HN dataset exhibited the smallest range of importance values, while most features were observed frequently. The RN and HP datasets showed a similar pattern, with the HP dataset being particularly noteworthy due to a subset of features displaying lower frequencies but higher importance values. The RP dataset displayed the largest number of features with high levels of both frequency and importance values.

Features were assigned weights, a combined metric of both relative frequency and importance, then ranked and grouped into rank groups. Features were then classified with respect to putative functions using the human metabolome database (HMDB, https://hmdb.ca). Lipids and lipid-like molecules were found to be widely distributed across rank groups while most other putatively annotated classes of metabolites were predominantly associated with lower rank features (Supplementary Fig. 5). The vast majority of the highly ranked features remain unannotated. Indeed, only ∼7% of the complete set of features identified in this study were associated with metabolite information from HMDB.

### 3.3 Evaluation of Classifier Performance

Prior to the evaluation of the classifier performance, a neural network based autoencoder was used to reduce the dimensionality of the datasets while preserving informative representation of the original (Supplementary Fig. 6). Using the compressed dataset, the ability of each of the five classifiers (RFC, SVC, ADA, KNN, LRC) to correctly identify cancer samples and non-cancer controls was independently evaluated using four metrics: 1) Positive predictive value (PPV, a.k.a. precision), 2) negative predictive value (NPV), 3) f1-score (F1), and 4) Matthew’s correlation coefficient (MCC). PPV (precision) is the number of true positives divided by the number of true positives plus false positives (potential range: 0-100%), while NPV is the number of true negatives divided by the number of true negatives plus false negatives (potential range: 0-100%). The f1-score, which symmetrically represents both precision and recall in a single metric, is the harmonic mean of precision and recall (a.k.a. sensitivity; potential range: 0-100%). MCC reflects the correlation between the observed and predicted binary classifications (potential range: −1 to +1). An MCC of +1 represents a perfect prediction, 0 no better than a random prediction and −1 indicates total disagreement between predictions and observations. MCC considers true and false positives and negatives and is generally regarded as a balanced measure of predictive accuracy even if the classes are of very different sizes [14]. The performance of each of the five classifiers and the consensus classifier based on repeated cross-validation is presented in Table 1.

**Table 1.** Performance evaluation metrics for individual and consensus classifier.

| Dataset | Classifier | PPV | NPV | F1 | MCC | Dataset | Classifier | PPV | NPV | F1 | MCC |
| --- | --- | --- | --- | --- | --- | --- | --- | --- | --- | --- | --- |
| HN | SVC | 97% | 88% | 95% | 0.86 | RN | Consensus | 97% | 90% | 95% | 0.86 |
|  | Consensus | 96% | 89% | 95% | 0.85 |  | SVC | 97% | 88% | 95% | 0.86 |
|  | KNN | 96% | 88% | 94% | 0.83 |  | LRC | 97% | 88% | 95% | 0.85 |
|  | LRC | 95% | 89% | 94% | 0.83 |  | ADA | 96% | 91% | 95% | 0.85 |
|  | ADA | 95% | 90% | 94% | 0.83 |  | KNN | 95% | 90% | 94% | 0.84 |
|  | RFC | 95% | 88% | 93% | 0.82 |  | RFC | 95% | 92% | 94% | 0.84 |
| HP | Consensus | 95% | 89% | 94% | 0.82 | RP | Consensus | 96% | 92% | 95% | 0.87 |
|  | SVC | 95% | 88% | 93% | 0.82 |  | SVC | 96% | 91% | 95% | 0.85 |
|  | KNN | 94% | 88% | 93% | 0.79 |  | KNN | 95% | 90% | 94% | 0.84 |
|  | LRC | 94% | 87% | 92% | 0.78 |  | LRC | 95% | 90% | 94% | 0.83 |
|  | ADA | 93% | 91% | 93% | 0.8 |  | ADA | 95% | 93% | 94% | 0.84 |
|  | RFC | 93% | 88% | 92% | 0.78 |  | RFC | 94% | 92% | 93% | 0.81 |

While the performance of the individual and consensus classifiers varied across different datasets, the differences were minor. The HP dataset displayed a slightly lower performance relative to the HN, RN, and RP datasets. However, the overall performance was consistently high across the four datasets (PPV ≥ 93%; NPV ≥ 87%; F1 ≥ 92%; MCC ≥ 0.78; Fig. 3A). The cumulative confusion matrix from the consensus classifier (Fig. 3B) is generally consistent with these results demonstrating a relatively low misclassification rate of false negatives (∼2 to 3%) and slightly higher rate of false positives (∼11 to17%).

**Fig. 3.**
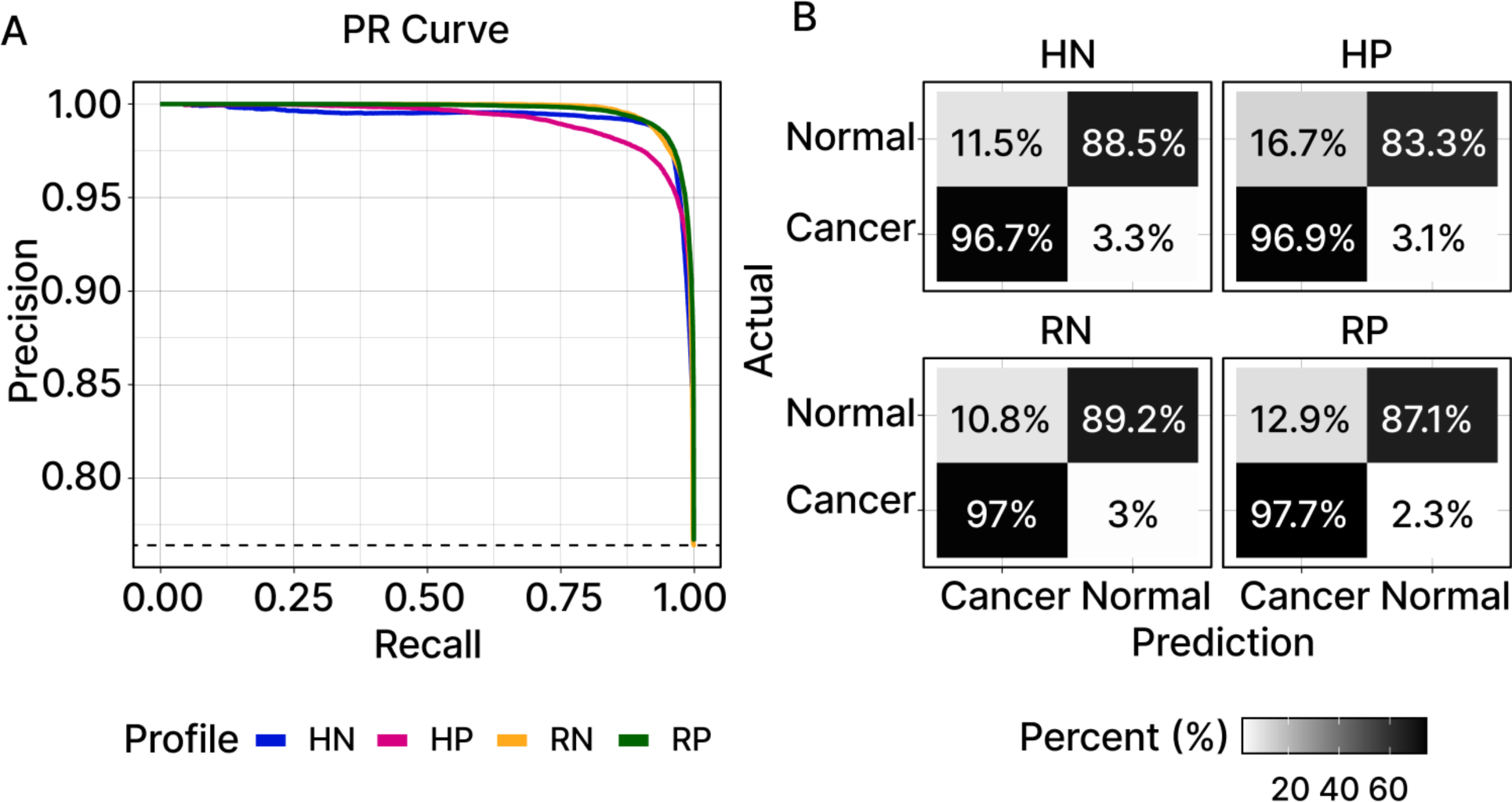
Comparison of consensus classifier performance. A) The performance characteristics of the models were graphically represented through precision-recall (PR) curves. Besides the HP dataset, the models for the remaining datasets showed similar levels of performance. B) Cumulative confusion matrices, also compiled from repeated CV, further reinforced these observations despite the false positives (FP) and false negatives (FN).

### 3.4 Utility of class probabilities as background distributions

The results of the repeated cross-validation scores can be used to assign a mean probability (adjusted to fall within a −2 to 2 range) that signifies the certainty of either a cancerous or non-cancerous classification. These probabilities were averaged for each sample and the distributions for each of the four datasets are displayed in Fig. 4A. The results highlight the classifier’s ability to clearly distinguish between cancerous and non-cancerous samples in all four datasets.

**Fig. 4.**
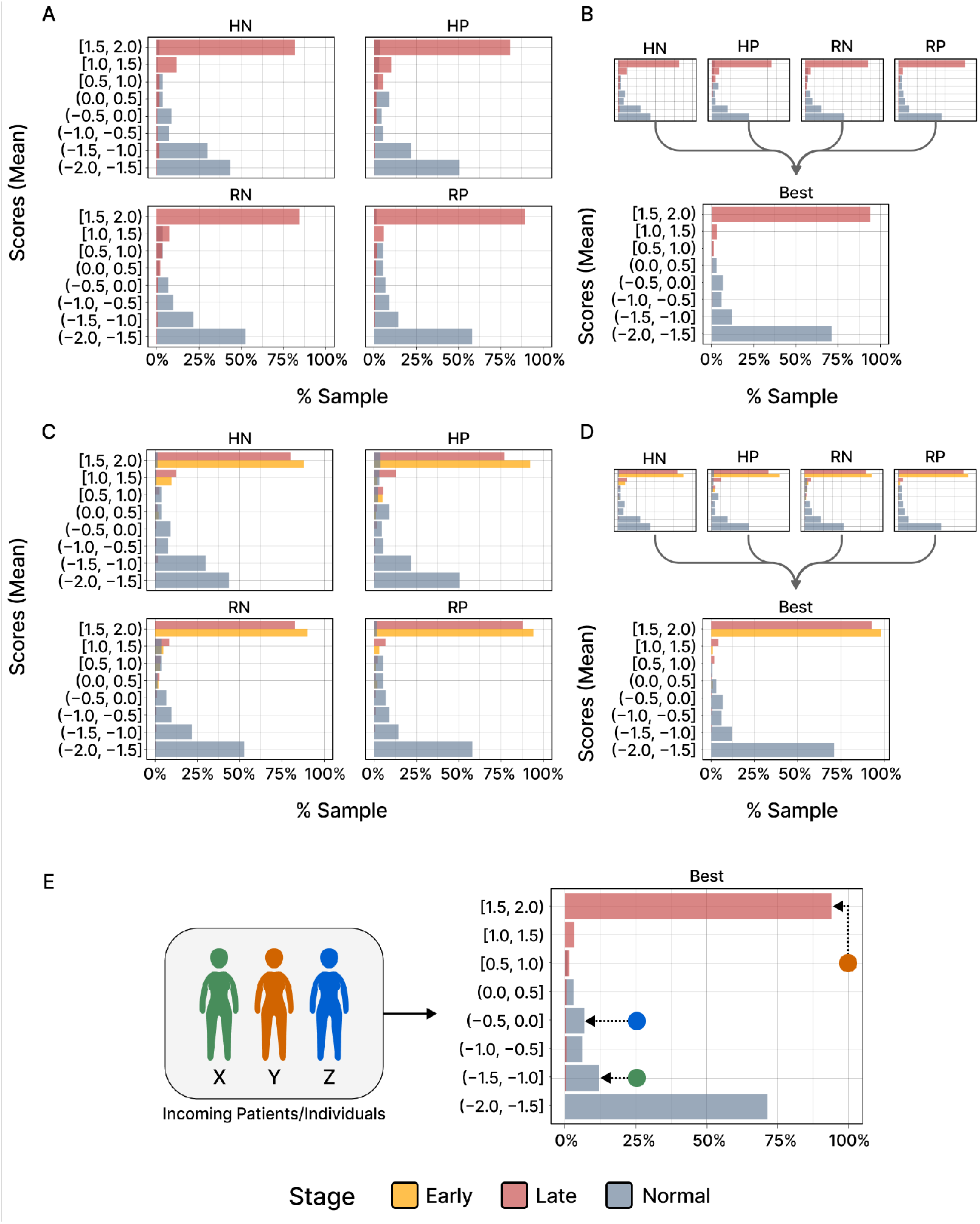
Evidence for the classifier’s ability to clearly distinguish between cancerous and non-cancerous samples. A) The bar charts exhibit the distributions of scores that have been converted from class probabilities and ranged from - 2 to 2 to improve visual clarity. The scores represent averages obtained from repeated cross-validation (CV) for each sample. Clear differentiation can be observed between the scores of cancer (red) and normal (blue) samples across all four datasets. The peaks in the distributions indicate the most frequently occurring score range for the samples. B) Similar to the previous bar chart, this figure illustrates the best average score across the four datasets demonstrating a notable improvement in classifying both cancer and normal classes. C) The bar chart illustrates the distribution of samples across various score ranges. This revealed that early- and late-stage samples clearly distinguish themselves from the normal samples. D) In an analogous manner to figure B), selecting the best score improves the final score for the scores at the stage-level. E) Diagram visualizing the potential adoption of the proposed workflow in a clinical setting. Given the absence of approved screening methods for ovarian cancer, this approach enables women to undergo serum profiling at a clinic to predict their cancer status. This could result in three possible scenarios: an individual’s serum profile (X) falls within a score range where misdiagnosis is unlikely, enabling a confident ruling out of a cancer diagnosis. An individual’s score (Y) falls within a range where 94% of others with this score have been diagnosed with cancer. Lastly, determining the cancer status of an individual (Z) may be challenging, as there are only a few samples within this score range and it is in the middle of the distribution.

By combining results from the four datasets and selecting the best average score among them, we observed a notable improvement in classifying both cancer and normal samples. This underscores that each dataset brings its unique contribution to the accurate prediction of cancer or non-cancer status (Fig 4B). A striking 97% of the cancer samples scored within the 1.0-2.0 range, with no (0%) misclassification of non-cancerous samples (Supplementary Table 7). In contrast, 83% of the non-cancerous/normal samples were found to fall within the −2.0 to −1.0 score range indicating that our consensus classifier is better at predicting cancer than non-cancer.

When binary classification results for the cancer samples are subdivided into early-stage (Stage I/II) and late-stage (Stage III/IV) cancer groups, the classifier still demonstrates high accuracy. It not only identifies late-stage cancer samples effectively but also classifies early-stage samples accurately. This holds true when considering both individual scores (Figure 4C) and best scores (Figure 4D). Using the best scores, the classifier’s predictive accuracy reaches 98% for early-stage samples and 92.7% for late-stage cancers for score range 1.5-2 (Supplementary Table 7). Adoption of our proposed workflow in a clinical setting would enable women to undergo serum profiling at a clinic to predict their cancer status (Fig. 4E).

## 4. Discussion

Although the incidence of OC is relatively low (2.5% of all malignancies in women [15]), it is among the most lethal of all cancers due to its high mortality rate. The reason for this is largely attributable to the fact that the disease is not typically diagnosed until the late (post-metastatic) stages of development (Stage III/IV) when effective treatment is difficult. For example, the most common sub-type of OC, serous papillary (65% of OC patients), is typically not diagnosed until Stage III/IV when the 5-year survival rate is only 31%. In contrast, if the disease is identified and treated early in its development (Stage I/II), the 5-year survival rate is 93%. These statistics dramatically underscore the dire need for an early diagnostic test for OC and other cancers where early-stage clinical symptoms are virtually non-existent.

The traditional approach for the identification of non-invasive biomarkers of cancer has been the screening of blood (or other body fluids) in search of significant changes in the presence/levels of molecules (typically proteins) associated with the disease [16]. A well-known example of such a diagnostic is the PSA (prostate specific antigen) biomarker for prostate cancer [17].

The OC biomarker candidate, CA125 (a.k.a., mucin 16/ MUC16) [18] was first introduced in 1996 [19]. Although an elevated level of CA125 is detected in ∼90% of late stage (III/IV) OC patients, it is elevated in only ∼50% of early-stage patients making it a poor biomarker of early-stage disease with a PPV of only ∼30% [20]. In 2003, a second candidate biomarker for OC, HE4 (human epididymis protein), was introduced [21]. While HE4 was an improvement over CA125 in having a reported PPV of ∼58%, it is still not sufficiently accurate to serve as a diagnostic test. Combining the results of the HE4 and CA125 together did not significantly improve PPV. However, the combination did lead to the development of a logistic regression model called ROMA (risk of malignancy algorithm) that was approved by FDA in 2011 as a method to classify patients with a pelvic mass into those with high vs. low risk of having OC [20].

By the early 2000s, it was becoming progressively clear that, on the molecular level at least, cancer was a much more complex disease than originally envisioned [4]. This realization was supported by findings indicating the existence of a multitude of disrupted molecular pathways (and underlying mutations) capable of leading to even the same cancer type. Such molecular level heterogeneity among individual cancer patients made the likelihood of identifying one or two biomarkers capable of accurately diagnosing all individuals with even the same type of cancer highly unlikely. As a consequence, the search for more accurate ways to diagnose cancer became focused on exploring larger combinations of biomarkers that might better capture the molecular heterogeneity underlying the disease [22]–[25].

In the case of OC, there were a number of multi-biomarker diagnostic tests developed in the early 2000s [26]–[28]. However, none of these early efforts were sufficiently validated to acquire FDA approval. In 2009, an assay (trade name OVA1) was proposed that incorporated levels of five serum proteins combined with proprietary software to generate high or low probability that an ovarian mass was a malignant tumor [29]. While the test was approved by FDA as a clinical aid in determining if a patient should be referred for further analysis, the test’s low PPV (31%) [30] eliminated it from consideration as an effective OC diagnostic. A more recent version of the OVA1 test (initially known as OVA2 but now trademarked as OVERA) uses a slightly different set of proteins upon which to generate its predictions. Although an improvement over OVA1, OVERA continues to be associated with a relatively low PPV (∼40%) [31], thereby again excluding it as a reliable OC diagnostic.

With the expanded availability of omics technologies and associated datasets (*e.g*., genomic, transcriptomic, proteomic, metabolomic) in recent years, a new approach to diagnostics began to emerge [32]. The application of various AI (artificial intelligence) approaches, most notably machine learning, to the analysis of large omics datasets of diseased and non-diseased individuals opened the possibility of the identification of patterns by which these categories could be distinguished. Predictive models built upon such classifications might then constitute a new generation of diagnostic tests.

While this basic concept is straightforward, its application is certainly not. There are multiple approaches to ML, and each is associated with individual strengths and weaknesses [11], [12]. In addition, the output from ML analyses of omics datasets is heavily dependent upon both the quality and type of data being analyzed. For example, classifiers that are based exclusively on ML analysis of DNA sequence datasets may be appropriate if the onset and progression of the disease in question is exclusively attributable to genetic mutations. Certainly, there is a significant genetic component to cancer, but other environmental (*e.g*., diet, lifestyle, microbiome, *etc*.) and molecular (*e.g*., epigenetic/gene expression changes, gene-gene/protein-protein interactions, *etc*.) factors are also known to play significant roles. Indeed, it has been proposed that the accurate characterization of a complex disease like cancer will ultimately require simultaneous analyses of multi-omics datasets [33]. While this may well be the case, the development of computational methodologies sufficiently complex to accurately characterize multi-omics datasets is only in its infancy [34], [35].

In lieu of an approach that simultaneously analyzes multi-omics datasets, we chose a currently available alternative, *i.e*., working with a dataset that reflects biological changes occurring on multiple levels. Metabolic profiles are widely viewed as a molecular phenotype reflective of underlying collective information encoded at the genome level and realized at the transcriptome and proteome levels. As such, metabolic profiles have long been considered promising indicators of cancer and other complex diseases [8], [9].

To help ensure the quality of our metabolic data, individual normal and OC patient samples were collected from four geographically divergent locations and analyzed using ultra-performance liquid chromatography coupled with tandem mass spectrometry (UPLC-MS/MS-positive and negative modes and each sample independently pre-processed through two columns) generating four distinct datasets (HN: HILIC negative; HP: HILIC positive; RN: C_18_ reversed phase negative; RP: C_18_ reversed phase positive). To guard against instrumental drift between runs, the same control samples were analyzed following every ten biological samples. Principle component analyses of data generated from our MS analyses across different batches and times demonstrated little experimental variation between runs.

Each of our four datasets was analyzed separately to determine if any particular dataset contained more relevant information than any other. We found little difference in the accuracy of predictions computed using each dataset individually. When we combined the best average score from each of the four datasets, we observed an improvement in the classification of both cancer and normal samples. This suggests that each of the four datasets contribute uniquely to the accurate prediction of cancer status.

Computationally, we evaluated the performance of five independent ML classifiers. A consensus classifier that generates average predictive probabilities from the probabilities of each of the individual classifiers gave the best overall performance with a PPV of 93%. Interestingly, the overall predictive accuracy of our consensus classifier was better for early-relative to late-stage patients. We found that late-stage patients display greater heterogeneity in molecular profiles than early-stage patients. While the reason for this dichotomy is currently unknown, the preliminary findings suggest that OCs may become more metabolically heterogeneous as they progress/metastasize. However, because the sample size of early-stage patients is considerably less than late-stage patients in this study, further analysis of expanded datasets will be required to resolve this issue.

Our model’s accuracy in predicting women with OC is slightly greater than its accuracy in predicting women without the disease. The reason for this is currently unknown but may, at least in part, be due to the fact that the model may be detecting disease in women prior to clinical symptoms and clinical diagnosis. Time course studies are currently being instituted to test this hypothesis.

The high PPV (93%) associated with our consensus classifier supports the notion that ML analysis of omics data, and of metabolomic data in particular, is an extremely promising approach for the future diagnosis of ovarian and possibly other cancers as well. Such analyses will likely lead to a more probabilistic approach to cancer diagnosis that will serve to personalize the process much as genomic profiling of individual patient tumors is personalizing cancer treatment (*i.e*., precision cancer medicine).

Despite these highly favorable prospects, it is important to keep in mind the limitations of ML analyses of omics data. For example, the PPV associated with even the same ML based predictive model can be highly sensitive to the size and composition of the datasets employed in building and testing the models. For example, in an earlier pilot study of the metabolic profiles of a relatively small number (46) of OC patient samples collected from one of the same areas sampled in our current study (Northside Hospital, Atlanta), the authors generated a predictive model with a putative accuracy of 100% [36]. The relative reduction in accuracy associated with our current model relative to this earlier study coupled with the fact that none of the top ranked features in the earlier study ranked within the top 100 features in our current study (Supplementary Table 8) underscores the impact of datasets on ML/metabolomic based predictive models. Future refinements in the development of metabolomic (and likely all omics) based ML models will need to address the issue of how many samples over what geographic area are needed to reflect the full spectrum of diversity in OC (and other cancer types).

In an effort to exemplify how the type of results generated in our study might, in the future, translate into a clinically useful tool, we grouped the quantity and percentage of our samples into score ranges. We envision a clinical tool in which the scores of individual patients can be mapped across such a distribution providing a likelihood that an individual patient does or does not have cancer. Such information could serve as a significant aid in determining the need for treatment or continued monitoring. For example, consider the scenarios presented in Fig. 4F. Scenario (A) represents a situation in which an individual’s serum profile falls within a score range that makes cancer highly unlikely. In such a case, the individual may only require yearly monitoring. In scenario (B), the detection is more problematic due to a relatively small number of samples in this score range and a comparable number of cancerous and non-cancerous patients. In such cases, a patient may be referred for more additional and/or more frequent screening. Scenario (C) depicts a situation where an individual’s score lies in a range where a majority (94%) of patients has been diagnosed with cancer. In such a case, the patient would likely be referred for immediate advanced screening/treatment.

In summary, our results confirm the overall potential of an integrative approach using metabolomic profiles and ML-based classifiers for the detection of OC. The accuracy of these classifiers is highly dependent upon both the quality and quantity of the data upon which models are built. We found little difference in the accuracy of predictions generated using alternative ML classifiers, although the consensus classifier generated the most accurate predictions. Application of results generated from our consensus classifier illustrated how the frequency distribution of individual patient scores can be used to develop a useful clinical tool that assigns a likelihood that an individual does or does not have OC. We believe this personalized/probabilistic approach to cancer diagnostics is more robust and clinically informative than the more traditional binary (yes/no) tests and may represent a promising new direction in the early detection of OC and perhaps other cancer types as well.

## CRedit authorship contribution statement

**Dongjo Ban:** Conceptualization, Formal analysis, Investigation, Methodology, Software, Roles/Writing-original draft. **Stephen N. Housley:** Formal analysis, Software, Writing - review & editing. **Lilya V. Matyunina:** Data curation. **L. DeEtte McDonald:** Data curation, Writing - review & editing. **Victoria L. Bae-Jump:** Data curation. **Benedict B. Benigno:** Data curation, Funding acquisition. **Jeffrey Skolnick:** Formal analysis, Writing - review & editing. **John F. McDonald:** Conceptualization, Investigation, Project administration, Resources, Supervision, Roles/Writing -original draft, Writing - review & editing.

## Data Availability

All data produced in the present work are contained in the manuscript

## Acknowledgements

We thank Ramit Bharanikumar and Zainab Arshad for their contributions to pilot studies related to this project.

## Supplementary Material

Supplementary material to this article can be found online at https://

