## supplemental material for "A Personalized Probabilistic Approach to Ovarian Cancer Diagnostics"

**Methods in detail**

*Sample collection*

Serum samples from ovarian cancer (serous papillary) patients and non-cancer controls were obtained from: 1) Northside Hospital, Atlanta, GA (10 early- and 142 late-stage cancer samples); 2) Fox Chase Cancer Center Biosample Repository Facility, Philadelphia, PA (51 early- and 68 late-stage cancer samples, 133 control samples); 3) University of North Carolina Medical School, Chapel Hill, NC (17 early-stage cancer samples); 4) Alberta Health Services, Alberta, Canada (23 early- and 120 late-stage cancer samples). Serum samples were aliquoted (500ul) into separate tubes and stored at -80 °C.

*Sample Preparation*

Serum samples were sent to Creative Proteomics (New York) for analysis. 100 µL of each serum sample were transferred into a 1.5 mL Eppendorf centrifuge tube with 300 µL of 80% methanol added into each tube before the mixture was vortexed for 30 seconds. Subsequently, the samples were kept at -40 °C for 1 hour. Samples were vortexed again for 30 seconds and then stored once more at -40 °C, for 30 minutes. Following this, the samples were centrifuged at 12,000 rpm and 4 °C for 15 minutes. The supernatant was carefully transferred to clean tubes and kept at -40 °C for an additional hour. After this last incubation, the samples were centrifuged again under the same conditions for another 15 minutes. Finally, 120 µL of the supernatant and 3 µL of DL-o-Chlorophenylalanine (100 µg/mL concentration) were transferred to a vial, ready for UPLC-MS/MS analysis. In addition to the individual patient samples, pooled quality control (QC) samples were created by combining small aliquots of the study samples in order to monitor the performance and consistency of the analysis.

*UPLC-MS/MS Analysis of Serum Samples*

Separation of the molecules was performed using C18 (reversed phase, "R") and HILIC (hydrophilic interaction liquid chromatography "H") columns with the UltiMate™ 3000LC combined with Q Exactive MS (ThermoFisher). For C18, the LC system includes an ACQUITY UPLC BEH $C_{18}$ (100×2.1mm×1.7 µm). The mobile phase contains solvent A (0.05% formic acid-water) and solvent B (acetonitrile) with a gradient elution (0-1.0min, 95% A; 1.0-12.0 min, 95%-5% A; 12.0-13.5 min, 5% A; 13.5-13.6 min, 5%-95% A, 13.6-16 min, 95% A). The flow rate of the mobile phase was 0.3 mL/min. For HILIC, the LC system is comprised of XBridge BEH Amide Column (150×4.6 mm×3.5 µm) with mobile phase utilizing solvent A (5% acetonitrile water, containing 10 mM ammonium formate) and solvent B (95% acetonitrile-water, containing 10 mM ammonium formate) with a gradient elution (0-0.5 min, 5% B; 0.5-9.5 min, 5%-35% B; 9.5-11.5 min, 35%-60% B; 11.5-13.5 min, 60% B; 13.5-13.6 min, 60%-5% B; 13.6-16.0 min, 5% B). The flow rate of the mobile phase was 0.6mL/min. The temperature for both columns was maintained at 40 °C with the sample manager temperature set at 4 °C. For every analysis, the injection volume of each sample was 10 µL with a QC sample being analyzed every ten patient samples.

The following mass spectrometry (MS) parameters were used during positive ("P") and negative ("N") ionization modes using electrospray ionization (ESI): heater temperature of 300 °C, sheath gas flow rate of 45 arbitrary units (arb), aux gas flow rate of 15 arb, sweep gas flow rate of 1 arb, and capillary temperature of 350 °C. The spray voltage and S-Lens RF level for ESI+ and ESI- modes are 3.0KV and 30%, and 3.2KV and 60%, respectively.

*Data generation*

Two different MS data acquisition methods were used during this study: full scan and data-dependent scanning (ddMS2). For full scan data acquisition, the resolution was set at 70,000 full width half max (fwhm), the automatic gain control (AGC) target was $3x10^6$, the maximum injection time was 100 ms, the scan range was

70-1050 mass-to-charge ratio (m/z), the polarity was set to negative or positive, and the spectrum data type was centroid. For ddMS2, the resolution was set at 17,500 fwhm, the AGC target was $1x10^5$, the maximum injection time was 50 ms, the isolation window was 1.7 m/z, loop count was 10, the normalized collision energy (NCE) for fragmentation of selected precursor ions based on the specified isolation window was 15/30/45 eV, and the spectrum data type was centroid.

The raw data from the scans consists of spectra that represent the intensity of the ions at different m/z where each peak in the spectra corresponds to a different ion detected. In the context of metabolomics analysis, these ions are referred to as "features", which represents the detected metabolites or analytes. Compound Discoverer Software (version 3.1) from ThermoFisher was used to process the raw data using the following parameters. For processing spectra data, minimum precursor mass was 0 dalton (Da), maximum precursor mass was 5,000 Da, and signal-to-noise threshold was 1.5. For the alignment model used to correct variations in retention times across different runs, adaptive curve was employed with maximum shift of 2 minutes and mass tolerance of 5 parts per million (ppm). Putative annotations of the features were assigned for preliminary identification from database searches using Compound Discoverer.

*Data Preprocessing*

The metabolomic datasets acquired through UPLC-MS/MS analysis of patient serum samples required multiple preprocessing stages to minimize inherent analytical errors. These errors are generally divided into two primary categories: systematic and random errors.

Systematic errors, also referred to as repeatable inaccuracies, are primarily introduced as batch effects and longitudinal drifts during sequential sample processing. To combat these, we employed a strategy known as Systematic Error Removal using Random Forest (SERRF) [1]. This approach leverages pooled QC samples to approximate and correct batch and injection order effects (Supplementary Fig. 1). To evaluate the effectiveness of this correction, non-parametric residual standard deviations (nRSDs) were calculated both pre- and post-correction.

Alongside systematic errors, data may also be influenced by random errors—unpredictable factors that are difficult to identify and quantify. Although these random errors present a unique challenge, it is possible to alleviate their impact by filtering features based on defined dispersion ratios (D-ratios) that measures the balance between technical and biological variation, with a lower D-ratio indicating higher quality and reliability. Filtering for D-ratio and nRSDs based on a given threshold ($< 0.1$ for this study) ensures the inclusion of only reliable and consistent features in downstream analyses (leaving only features that satisfy both nRSD and D-ratio thresholds). The number of features satisfying the said thresholds as well as the distribution of nRSDs and D-ratios can be found in Supplementary Table 1 and Supplementary Fig. 2, respectively.

*Workflow*

The workflow outlined in this study consists of three main steps. The first involves examining frequencies and importance values of the features in each of the four datasets using recursive elimination with cross-validation (RFECV). This specific task serves to identify stable "drivers" that would be important when making downstream predictions. To this end, a combined metric, "weight", is computed for each feature, allowing for rankings among the features within each dataset.

With these rankings, putatively annotated features were grouped based on their characteristics and functions. These rankings are taken into consideration during feature extraction via autoencoder (see below). This reduces the dimensionality of the original datasets prior to evaluating the consensus classifier that uses five individual classifiers to make predictions.

Finally, the probabilities assigned to individual samples in the study during the classifier evaluation step were further examined. This additional assessment serves not only to gauge the confidence of the predictions, but also to create a background distribution of the class (*Cancer/Normal*) probabilities that may potentially have a clinical utility in determining an individual's likelihood of having cancer. The workflow was specifically designed for binary classification, involving *Cancer* and *Normal* classes.

*Recursive Feature Elimination with Cross-validation (RFECV)*

As outlined above, RFECV was used to obtain information about the features in the dataset. This process involves selecting the most important features in a model, starting with the model training on all features. Then at each iteration, the least informative features are pruned from the current set of features until the algorithm reaches the specified number of features. Coupling RFE with CV ensures a more robust selection of the least informative features for removal. In evaluating the model during RFECV, the f1-score was used as the performance metric.

The RFECV implementation employed in this study utilized a tree-based ensemble method known as "extremely randomized trees" [2]. As with other ensemble methods in this category, it starts with multiple decision trees that are responsible for dividing the samples based on values of the features in the dataset. While other methods aim to choose the best feature and its value that can minimize the mix of *Cancer* and *Normal* classes after the split, the extremely randomized trees method randomly selects a feature and its value to perform the split. Each tree then computes a decrease in impurity using Gini index, reflective of the degree of heterogeneity resulting from the split.

The feature importance values, calculated by averaging the Gini index observed across all the trees in the ensemble, are utilized during RFECV to rank and prune features. This approach has demonstrated the ability to not only reduce over fitting but also maintain predictive performance. The implementation of RFECV, as described in this section, was done using ExtraTreesClassifier and RFECV with stratified K-fold cross-validation from scikit-learn library [3].

*Feature Assessment*

To assess the features of the datasets, RFECV was repeated 100 times with $K=5$ to examine both frequencies and mean importance values. Frequencies ($Fi$) refer to the number of times a given feature shows up throughout the iterations with the importance value ($GIi$) being the average Gini importance controlled for the frequency (Supplementary Fig. 3). These two metrics were merged using weighted average to compute weights for the features. Then, features were ranked using the weights of the features. In addition to giving the mean importance more relative influence in the final weight ($wi$) to be assigned, the decision of setting $\alpha$ to 0.3 was also influenced by its downstream impact on reconstruction error after the feature extraction step (Supplementary Fig. 4A) as well as the classifier evaluation (Supplementary Fig. 4B).

$$wi=\alpha * Fi+1-\alpha * GIi$$

Super class of a metabolite refers to a high-level categorization of compounds based on their chemical properties, functions, or biological activities. To assign such classes to the features, names and chemical formulas of the putative annotations were queried using the metabolite data (XML format) from Human Metabolome Database (HMDB) [4] to find a feature's super class (Supplementary Fig. 5, Supplementary Table 2).

*Feature Extraction Using Autoencoder*

An autoencoder is an artificial neural network often used for feature extraction where the goal is to learn a compressed representation of the original input data [5]–[7]. It consists of two components: an encoder and a decoder. The encoder outputs a lower-dimensional, or "latent", representation that captures the most important features of the input data. In contrast, the decoder evaluates the reconstruction error after decompressing the encoded data, providing a measure of how effectively the autoencoder preserved the original input. When implementing an autoencoder, there are components that require specification: the architecture of the autoencoder consisting of the number of layers and nodes per layer, activation functions, and optimizer.

In this study, an autoencoder with a single layer was implemented using the TensorFlow library [8]. This single layer, known as the bottleneck, performs compression to provide latent representation of the input data. The number of nodes in this layer was set to the square root of the number of features in the original data, which follows a heuristic that balances retaining information and dimensionality reduction [9], [10]. Increasing the number of nodes beyond this did not significantly improve the weighted mean squared error for the four datasets (Supplementary Fig. 6A).

Following the architecture setup, the exponential linear unit (ELU) [11] was selected as the activation functions for its ability to transform input values into a consistent range of real values post-compression. These functions are essential components of the autoencoder as they allow the autoencoder to learn and represent complex patterns found in the input data.

Finally, Adam optimization [12] was used to adjust the autoencoder's parameters with the goal of minimizing the difference between the autoencoder's reconstructed and the original data. By using this optimizer, the reconstruction process is improved over time by attempting to minimize the loss function, which quantifies the difference between the reconstructed ($Xi$) and the original data ($Xi$). Weighted mean squared error was employed as the loss function, which utilized the weights of the features ($wi$) computed during feature assessment to prioritize higher-ranked features during feature extraction.

$$Weighted\ MSE=1ni=1nwi{*}Xi-Xi2$$

In training the autoencoder, the default parameters from TensorFlow were used. These parameters include the batch size, the number of training epochs, and the learning rate.

*Consensus Classifier*

The implementation of consensus classifiers involves aggregating predictions from multiple classifiers to arrive at a final decision. Recognizing that different classifiers have different biases, strengths, and weaknesses, the consensus approach utilizes diversity to enhance its robustness [13].

In this study, five types of classifiers – logistic regression (LRC) [14], random forest (RFC) [9], support vector machine (SVM) [15], k-nearest neighbor (KNN) [16], and adaptive boosting (ADA) [17]– were employed using the implementations from scikit-learn library [3]. Hyperparameters for each classifier were fine-tuned using a grid search to ensure optimal performance prior to being incorporated into the consensus classifier (Supplementary Table 3). The probabilities generated from individual classifiers were calibrated using Platt scaling [18] to convert the output from individual classifiers into likelihood of prediction. Once ready, a soft-voting strategy was used to average the class probabilities predicted from each classifier to make the collective predictions.

To evaluate the performance of the individual and consensus classifiers in the presence of class imbalance, balanced accuracy, f1-score, average precision, and Matthew's correlation coefficient, sensitivity, specificity, negative predictive value (NPV), and positive predictive value (PPV) were used in conjunction with repeated cross-validation using stratified K-fold (*K=5*) strategy (Supplementary Tables 4,5). Moreover, cumulative confusion matrices were computed using the number of prediction outcomes collected from the repeated cross-validations. Finally, the performance of the consensus classifier was analyzed by shuffling the class labels to verify whether the classifier was truly learning meaningful patterns and relationships between the input features and original class labels rather than relying on random noise. The observed decrease in performance after shuffling the labels confirms the importance of using correct class labels during the classification (Supplementary Table 6).

The probabilities assigned to individuals in the study during cross-validation were utilized to create a background distribution of the class probabilities for *Cancer* and *Normal* classes. For visualization purposes, these probabilities were scaled to fit within a range of -2 to 2.

### Supplemental Figures

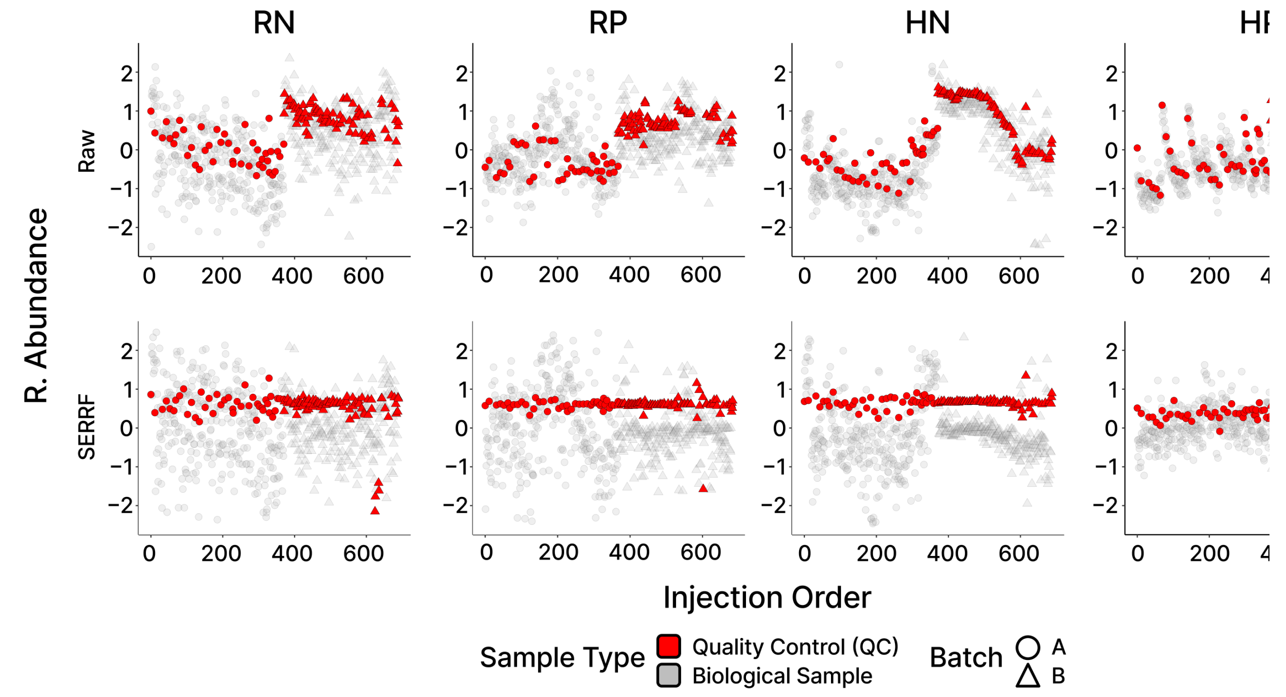


**Supplementary Fig. 1.** Relative abundance of select features before and after SERRF normalization. In the raw datasets, noticeable unwanted variation is observed between batches A and B in the QC samples, with significant drift within each batch. This drift can be attributed to instrumental drift and batch effects, which can introduce systematic errors. However, SERRF normalization effectively addresses most

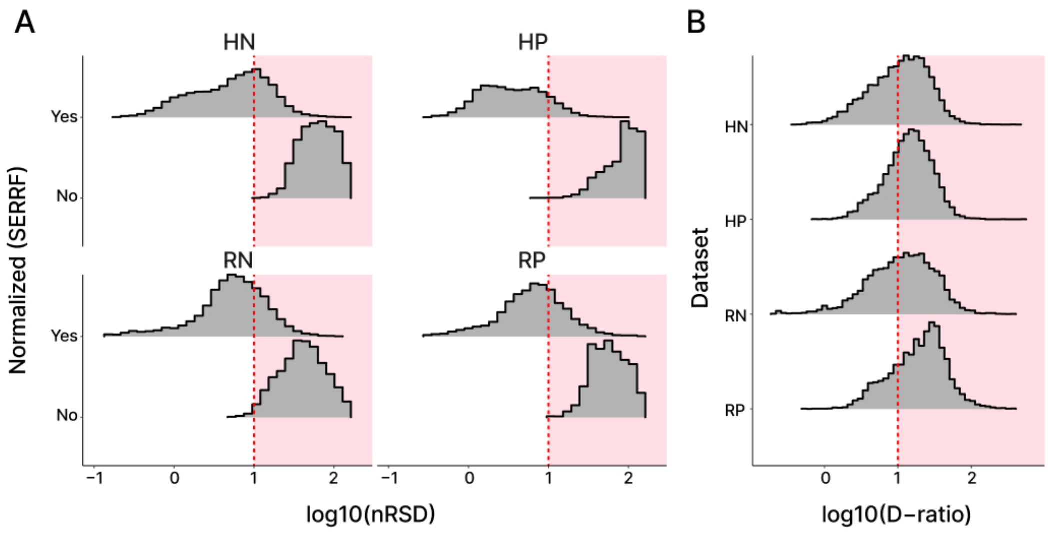


**Supplementary Fig. 2.** Feature quality control and filtering. A) Comparison of nonparametric residual standard deviation (nRSD) between raw and SERRF normalized datasets. Prior to applying the normalization, most features in the QC samples are observed in the red area (nRSD > 10%), which indicates high variability of the features. B) Distribution of dispersion ratios (D-ratios) observed in the normalized datasets. Features that are to be removed due to not meeting the threshold (D-ratio > 10%) are in the red area as they indicate higher variability in the QC samples than in the biological samples (R = C[illegible] reversed phase columns; H = HILIC columns;

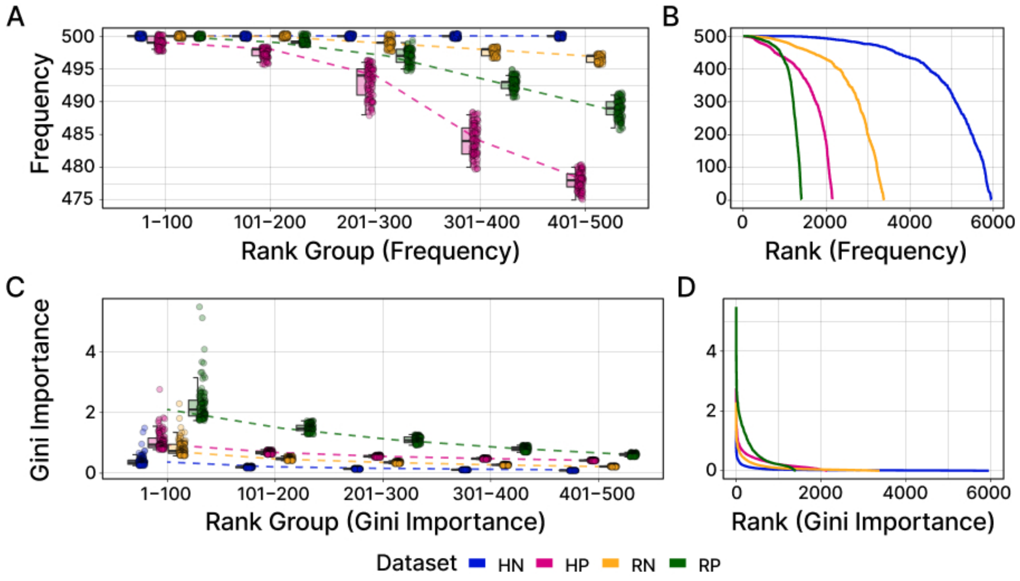


**Supplementary Fig. 3.** Frequencies and Gini importance values of the features obtained from RFECV. A-B) The frequencies plot demonstrates that, across the 4 datasets, a substantial proportion of the features are consistently observed across 100 iterations of RFECV (using stratified K-fold CV with ), rather than being limited to a select few features. C-D) The Gini importance values for the 4 datasets reveal a contrasting pattern. Features ranked below the top 100 highest Gini importance values exhibit relatively smaller importance values. Notably, in RP, a discernible pattern emerges with a presence of a few distinguishing features, while HN shows little difference between the importance of top-ranked features and those in the lower ranks.

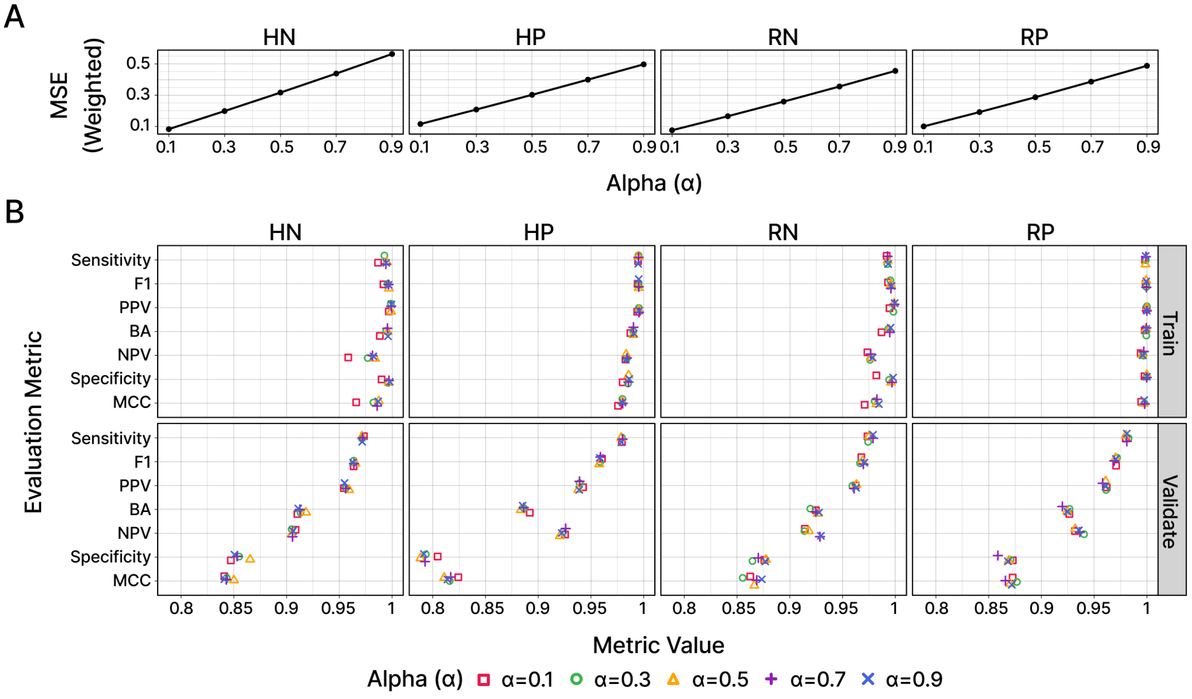


**Supplementary Fig. 4.** Effect of alpha on weighted mean squared error (MSE) and evaluation metrics. A) The line plot illustrates the relationship between alpha and weighted MSE. As alpha increases, the contribution of feature frequencies in the weight calculation also increases. As the alpha value increases, the reconstruction error from the autoencoder also increases. Therefore, prioritizing Gini importance appears more appropriate to retain the structure and information present in the original datasets. B) The scatter plot shows small differences in various evaluation metrics across different alpha values F1 = f1-score; PPV = positive predictive value; BA = balanced accuracy; NPV = negative predictive value; MCC = Matthew's correlation coefficient .

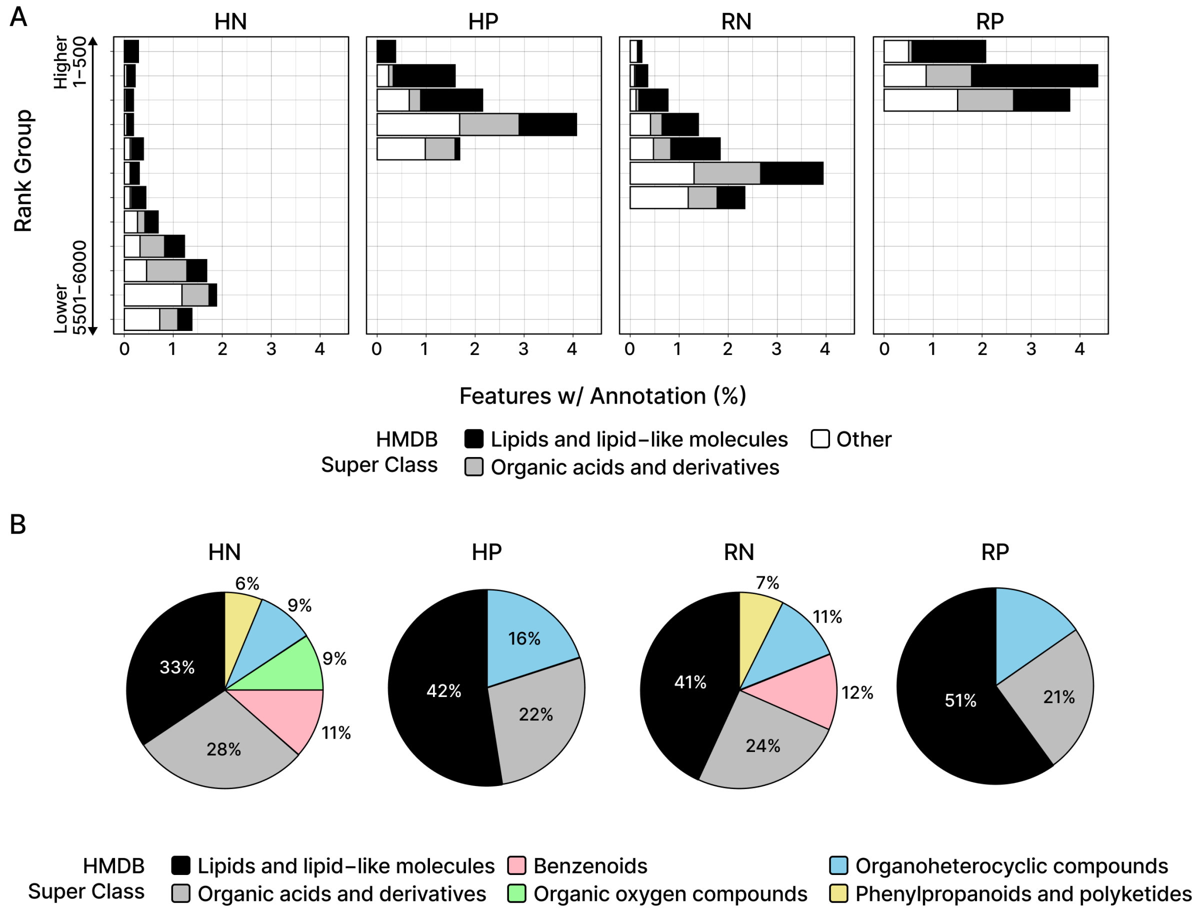


**Supplementary Fig. 5.** HMDB super classes of the putatively annotated features. A) The bar plot illustrates the percentage of features with putative annotations and their assigned HMDB super class. While a small percentage of the features have putative annotations, lipids and lipid-like molecules are the most observed super class compared to other super classes, particularly in higher ranks based on weights. B) The pie charts provide a more detailed examination of the putatively annotated features in terms of their classes. Consistent with the findings in A), lipids and lipid-like molecules are the most observed, followed by organic acids and derivatives. Other super classes are also displayed for comparison.

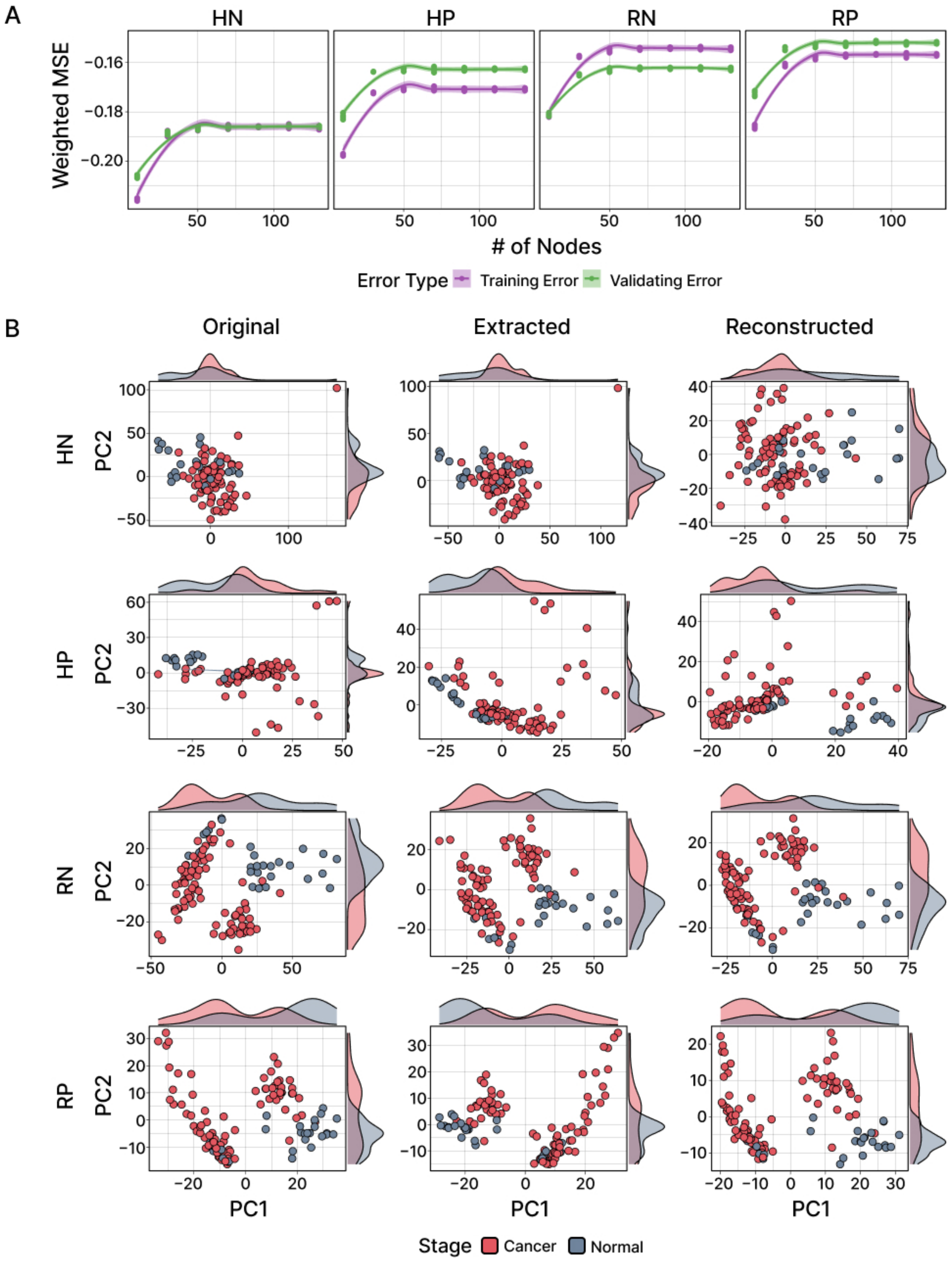


**Supplementary Fig. 6.** Assessment of feature extraction using autoencoder. A) Weighted MSE was examined for different number of nodes in the bottleneck layer during feature extraction. For all four datasets, the curve plateaus around 50 nodes, indicating that adding more nodes beyond this point adds no substantial improvement during reconstruction. B) Principal component analysis (PCA) was performed on the original input data, extracted features from the autoencoder, and reconstructed data from the extracted features. The scatter plot illustrates the representation of cancer and normal samples in the first two principal components from PCA. While the positions of the points change in the extracted and reconstructed data, the preservation of the original structure is evident from the density plots shown at the top and right sides of the scatterplots, where the distribution of both cancer and normal samples remains similar.
